# High-gamma and beta bursts in the Left Supramarginal Gyrus can accurately differentiate verbal memory states and performance

**DOI:** 10.1101/2024.05.29.24308117

**Authors:** Nicolás Sawczuk, Daniel Y. Rubinstein, Michael R. Sperling, Katrina Wendel-Mitoraj, Petar Djuric, Diego F. Slezak, Juan Kamienkowski, Shennan A. Weiss

## Abstract

The left supramarginal gyrus (LSMG) may mediate attention to memory, and gauge memory state and performance. We performed a secondary analysis of 142 verbal delayed free recall experiments, in patients with medically-refractory epilepsy with electrode contacts implanted in the LSMG. In 14 of 142 experiments (in 14 of 113 patients), the cross-validated convolutional neural networks (CNNs) that used 1-dimensional(1-D) pairs of convolved high-gamma and beta tensors, derived from the LSMG recordings, could label recalled words with an area under the receiver operating curve (AUROC) of greater than 60% [range: 60-90%]. These 14 patients were distinguished by: 1) higher amplitudes of high-gamma bursts; 2) distinct electrode placement within the LSMG; and 3) superior performance compared with a CNN that used a 1-D tensor of the broadband recordings in the LSMG. In a pilot study of 7 of these patients, we also cross-validated CNNs using paired 1-D convolved high-gamma and beta tensors, from the LSMG, to: a) distinguish word encoding epochs from free recall epochs [AUC 0.6-1]; and distinguish better performance from poor performance during delayed free recall [AUC 0.5-0.86]. These experiments show that bursts of high-gamma and beta generated in the LSMG are biomarkers of verbal memory state and performance.

## Introduction

The role of invasive technologies in treating the 15-20% of the population over 65 years old with mild cognitive impairment (MCI) is not yet established. Closed-loop stimulation or biofeedback could be utilized to improve attention and memory by detecting and improving poor memory encoding or recall states^1^. Herein, we show proof of concept that accurate neurophysiological biomarkers of memory states and performance can be derived exclusively from intracranial EEG (iEEG) recordings from the Left Supramarginal Gyrus (LSMG-Brodmann area 40). The LSMG is hypothesized to be important in directing attentional resources towards relevant external and internal (*i*.*e*., mental) stimuli for promoting memory encoding and retrieval^2–6^, and is part of the dorsal language network^7^ which is analogous to the right hemispheric ventral attention network^8^. An alternative hypothesis of LSMG function is that of a mnemonic accumulator in which LSMG activity correlates with memory state and performance but does not encode memory engrams^2–6^.

Past work utilizing averaged fMRI^4^ and iEEG signals^9^ show that, for subsequently recalled verbal and visual information, the LSMG signals show subsequent memory effects (SMEs). A SME is when the average recalled memory items’ physiological response, as compared to the average forgotten items’ response, is significantly larger (*i*.*e*., positive) or smaller (*i*.*e*., negative). During encoding, BOLD signal in the LSMG exhibits SMEs that can be either positive or negative^4^, and high-gamma (HG) power in the LSMG also exhibits a positive SME^9^. During stimulus recall epochs, BOLD signals^2,3,5^and HG power^10^ also exhibit a positive SME in the LSMG.

Since BOLD and iEEG activity in LSMG appears to correlate with memory, we examined 142 verbal delayed free recall experiments performed by medically-refractory focal epilepsy patients undergoing pre-surgical evaluation with depth and subdural electrodes placed, in part, in the LSMG (Figure 1A,^1,11,12^). The electrode contact placement was selected to best delineate the seizure onset zone and was unrelated to this secondary analysis of the verbal memory task. In the delayed free recall task, participants were instructed to study lists of words, and after an arithmetic distractor epoch, recall as many words as possible (Figure 1A1-3). The free recall task is often utilized to study the encoding and retrieval of episodically formed associations^13^.

**Figure 1:**
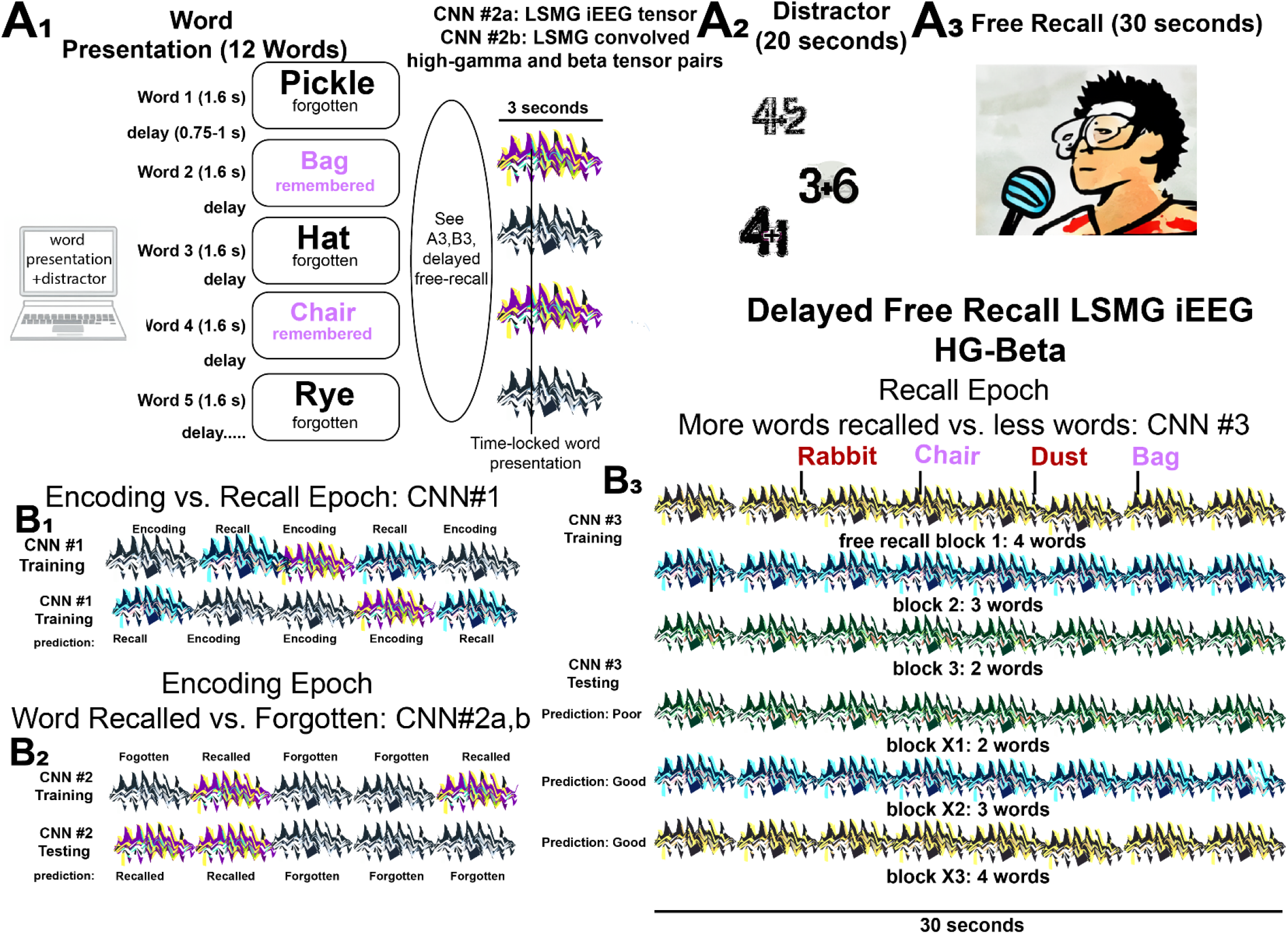
Experimental design for classifying performance in the delayed verbal free recall task using iEEG tensors and convolved high-gamma and beta tensor pairs to train and cross-validate convolutional neural networks (CNNs) A) The patients participated in a verbal delayed free recall task (encoding, distractor, and recall). First, the patient was instructed to remember a list of 12 words that were sequentially presented on a computer screen separated by an inter-word interval (A1). The illustration of LSMG iEEG tensors adjacent to the words represent either: 1) a one dimensional (1-D) tensor of the broadband LSMG iEEG recorded time locked to word presentation; or 2) a pair of 1-D tensors of LSMG convolved high-gamma (HG) and beta that are also time locked with word presentation. Whether the words, and their respective tensors, correspond with successful or failed word encoding is defined by the delayed free recall epoch (A3), that follows the arithmetic distractor epoch (A2). In the experimental block shown the patient successfully encodes two words “Bag” and “Chair” (magenta). (B) Illustrations of the training and cross-validated testing of the 3 CNNs using the pairs of (1-D) color-coded tensors of convolved high-gamma (HG) and beta burst timeseries recorded solely from the left supramarginal gyrus (LSMG) intracranial EEG (iEEG). Different colors are associated with the different tasks and the task performance (Key: black and white iEEG: encoding - forgotten; purple and yellow: encoding - recalled; black and green: recall - poor; black and blue: recall - good; black and green: recall - good). Note that CNN2a is distinct from CNN1,2b,3 because it utilizes the broadband iEEG as the 1-D tensor. Additionally, the recall epoch (B3) is not time locked to the spoken words and is evaluated solely by the number of words recalled correctly in the 30 second duration. In B1, the 3 sec. word-presentation encoding epochs and compared with random 3 sec. epochs from the free recall epoch.

We analyzed iEEG recorded from depth or sub-dural electrode contacts placed in, or on, the LSMG. We utilized iEEG time locked to specific task epochs (Figure 1) to derive, from the iEEG, pairs of one-dimensional (1-D) tensors consisting of: a) convolved HG bursts; and b) convolved beta bursts. These tensor pairs were then used to train and cross-validate convolutional neural networks (CNNs) that could differentiate verbal episodic memory state (i.*e*., encoding vs. recall, Figure B1) and performance during encoding (i.e., successfully encoded word, vs. failed encoding [forgotten]), Figure B2) or performance during recall (more words freely recalled during a session as compared to other sessions, Figure B3).

We hypothesized that HG and beta bursts in the LSMG would serve as adequate biomarkers of verbal encoding and recall in the verbal delayed free recall task because: 1) the LSMG has a critical role in mnemonic function^2–6^; 2) HG is strongly associated with cognitive performance, particularly when phase-amplitude coupled with theta oscillations^14^, and HG bursts are mostly attributable to pyramidal neuron firing and correlate with the BOLD signal^14–17^; and 3) gamma and beta bursts are generated by ensembles of inhibitory interneurons that coordinate the activity of excitatory pyramidal cells which is implicated in mediating working memory, and perhaps encoding individual working memory items in the frontal lobe^18–20^.

## Methods

### Participants

Patient data were collected as part of a multi-center project named Restoring Active Memory (RAM, Principal Investigator: Michael Kahana) https://memory.psych.upenn.edu/Main_Page#Cognitive_Neuromodulation at the following centers: Thomas Jefferson University Hospital (Philadelphia, PA), University of Texas Southwestern Medical Center (Dallas, TX), Emory University Hospital (Atlanta, GA), Dartmouth-Hitchcock Medical Center (Lebanon, NH), Hospital of the University of Pennsylvania (Philadelphia, PA), and Mayo Clinic (Rochester, MN). The research protocol was approved by the Institutional Review Board at each hospital and informed consent was obtained from each participant. A subset of this data, previously analyzed in (Rubinstein et al., 2021) and R01 proposal R01MH120161, was utilized by Dr. Weiss and colleagues in accordance with an agreement with Thomas Jefferson University. This data is also available at

FR1: https://openneuro.org/datasets/ds004789/versions/3.1.0

catFR1: https://openneuro.org/datasets/ds004809/versions/2.2.0.

For details on the depth and subdural electrodes used, the detailed electrophysiological data acquisition, and the neuroanatomical localization of the electrode contacts see^1,11,12^. We included patients who had as few as two intracranial EEG contacts in the LSMG, other RAM patients were excluded. Data were recorded with a minimum sampling rate of 500 Hz. Additionally, iEEG recordings from the LSMG were visually inspected for quality, and contaminated and noisy contact recordings were excluded prior to analysis with the convolutional neural networks. Recordings in the dataset that utilized electrical stimulation of the brain were excluded as well.

### Verbal memory task

Each patient participated in a delayed free-recall (FR) task. An experimental session consisted of encoding, distractor, and recall epochs. The encoding epoch consisted of 12 words, with each word shown on the screen for 1,600□ms, followed by a blank inter-stimulus interval with a duration between 750–1,000□ms. Immediately following the final 12^th^ word in each list, a distractor task consisting of arithmetic problems was performed for 20 s. Subsequently in the recall epoch, participants were then given 30□s to verbally recall as many words as possible from the list in any order (Figure 1A). Further details can be found in^1,11^. List words in some experiments were categorically organized (catFR). Patients completed between 8-50 sessions.

### High-gamma (HG) and beta burst amplitude analysis

All experimental sessions were visually analyzed in Micromed™ Brain Quick™. Consistency refers to the recurrence of these bursts during the encoding epoch. For measuring consistent peak HG amplitude, finite impulse response (FIR) band-pass filters between 80-200 Hz were used, and the peak amplitude was evaluated using the standard cursor across the recording duration. To measure consistent peak beta amplitude the FIR band-pass filters between 16-30 Hz were used instead, followed by evaluation using the standard cursor across the recording duration. The actual peak values were subtracted from the baseline values between the bursts.

### High-gamma (HG) and beta burst analysis

All custom code was written in Python™. The signal from each contact was standardized using StandardScaler (scikit-learn). The topographical analysis of the wavelet convolution^21^was adapted to detect the discrete HG and beta oscillatory bursts in the computed wavelet convolutions of the recording from each contact (Figure 2,3, Supplementary Methods). The topographical analysis of the wavelet convolution was utilized twice in frequency ranges overlapping with HG [50,250 Hz] and the beta bands [5,50 Hz] to define the onset time, offset time, duration, spectral content, and power of distinct bursts in both the HG and beta band. Rather than utilizing the raw band-pass filtered iEEG as pairs of 1-D tensors to train CNNs, the oscillatory bursts were represented with Gaussian kernel functions:

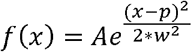

where A is the amplitude of the Gaussian scaled by the power of each oscillatory burst, p the midpoint between onset and offset time and w the time duration of each oscillatory burst.

**Figure 2:**
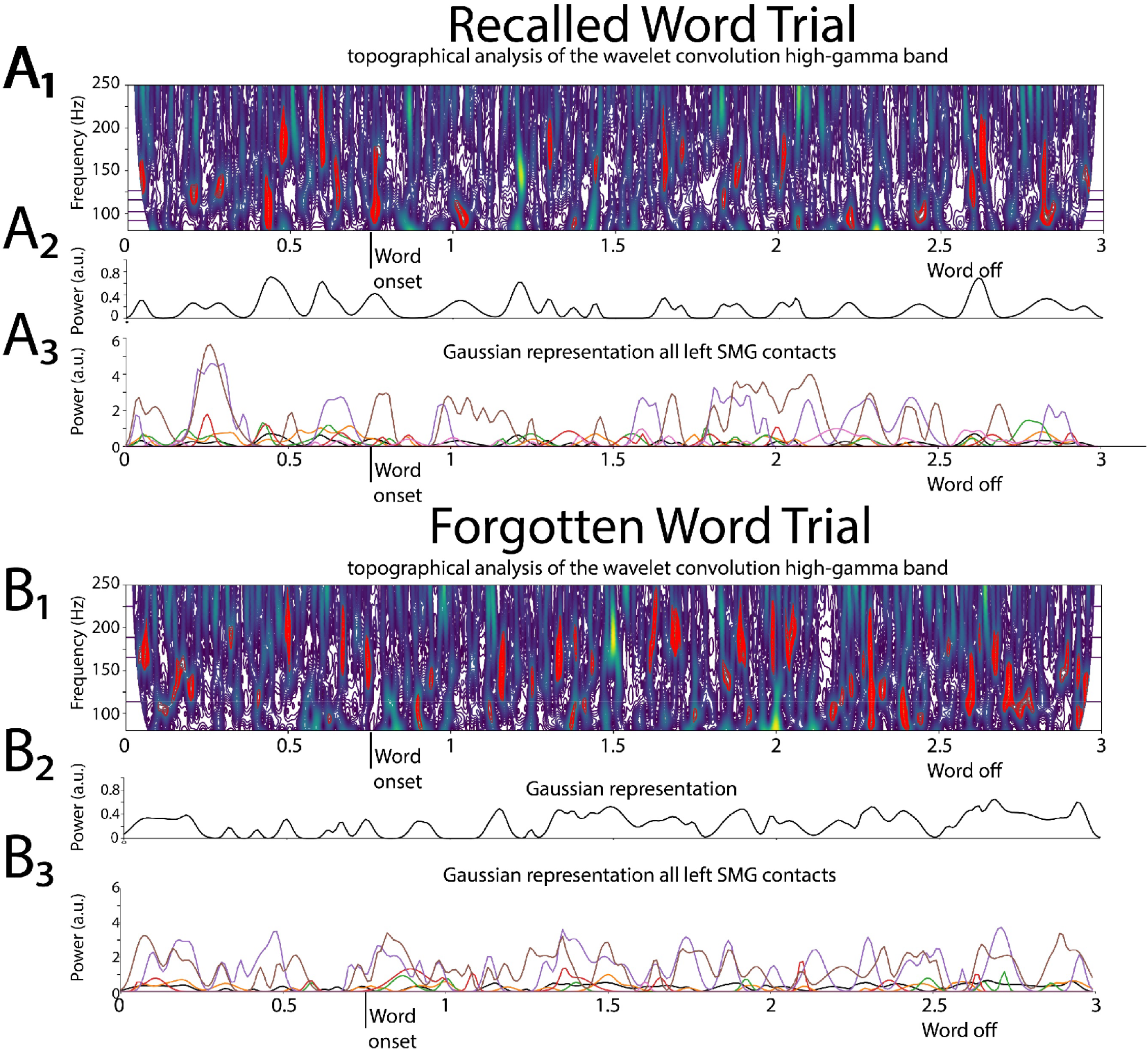
Illustrative example of deriving the 1-D tensor of convolutional high-gamma (HG) in the left supramarginal gyrus (LSMG): Shown in A1 and B1 are topographical analyses of the wavelet convolution, in the HG band, during stimulus-locked successful (A1) and failed (B1) word encoding trials. A2 and B2 are the respective Gaussian convolved time series derived from A1 and A2, respectively. Shown in A3 and B3 are the Gaussian convolved time series recorded by other contacts in the LSMG used to derive the 1-dimensional tensor of convolved HG, for a successfully encoded (A) and forgotten (B) word encoding trial.

**Figure 3:**
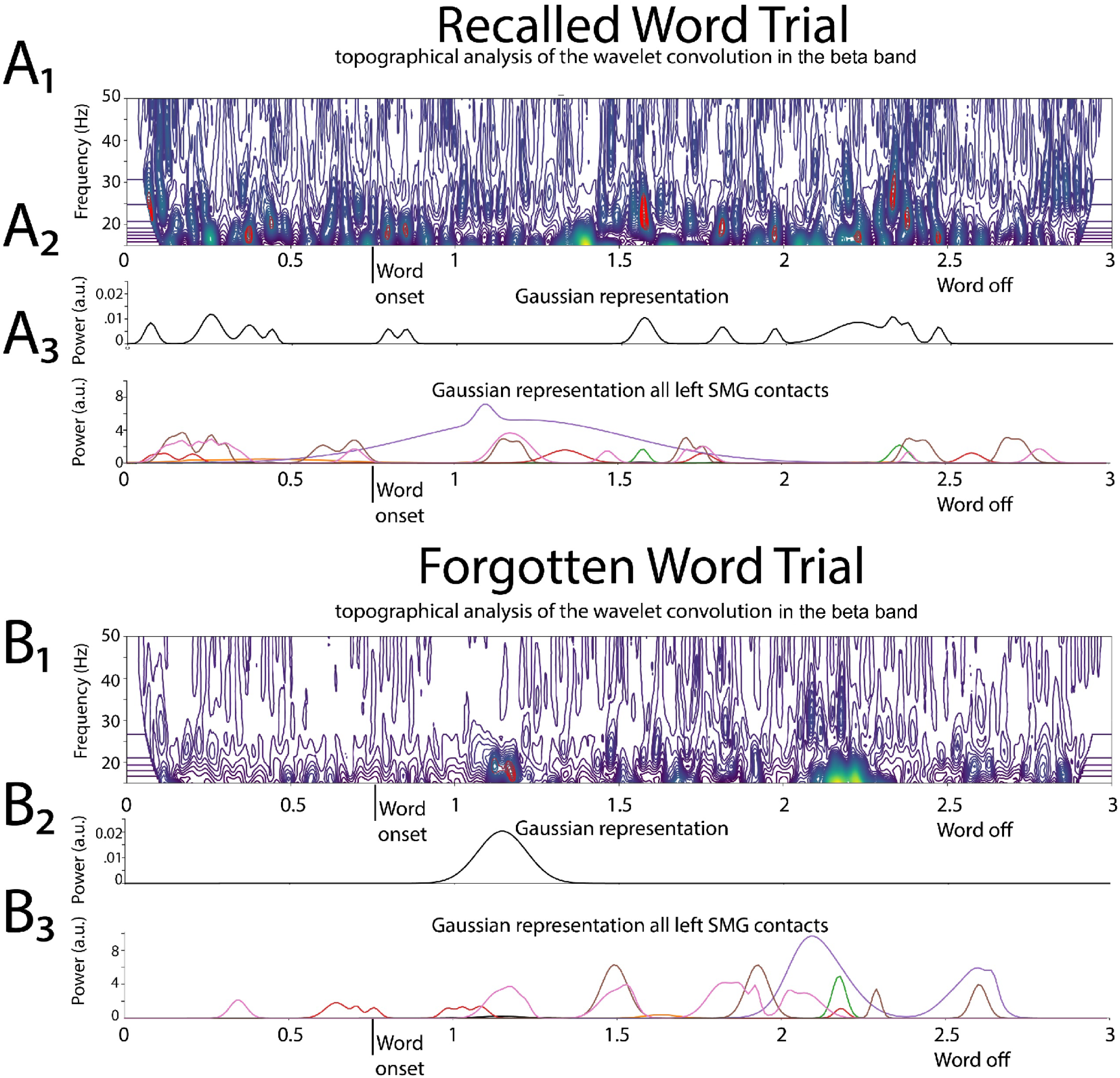
Illustrative example of deriving the 1-D tensor of convolutional beta in the left supramarginal gyrus (LSMG): Shown in A1 and B1 are topographical analyses of the wavelet convolution, in the beta band, during stimulus-locked successful (A1) and failed (B1) word encoding trials. A2 and B2 are the respective Gaussian convolved time series derived from A1 and A2, respectively. Shown in A3 and B3 are the Gaussian convolved time series recorded by other contacts in the LSMG used to derive the 1-dimensional tensor of convolved beta, for a successfully encoded (A) and forgotten (B) word encoding trial.

For a single LSMG iEEG recording from a single contact during a specific epoch of the task, the burst analysis generates both a single HG burst Gaussian (*i*.*e*., convolved) timeseries and a single beta burst Gaussian (*i*.*e*., convolved) timeseries. Since all experiments utilized more than one recording contact in the LSMG, a pair of 1-D tensors were generated for bursts of: 1) the convolved HG; and 2) the convolved beta (Figure 2, 3).

### Convolutional Neural Network (CNN) Training and Cross-Validation

To train the CNN that can differentiate encoding from recall memory states (Fig 1B1, CNN1), the pairs of 1-D convolved HG and beta tensors, derived from the LSMG iEEG during the 3-second stimulus locked word encoding trials, were combined at random with 3-second pairs of 1-D convolved HG and beta tensors randomly selected from the LSMG iEEG during the 30 second verbal free recall epoch. The CNN1 was then trained using the concatenated and correctly labeled randomized paired 1-D convolved HG tensor and convolved beta tensors.

CNN2a and 2b were trained to differentiate between recalled and forgotten words during the stimulus locked word encoding epoch iEEG recordings (Fig 1A2,B2). CNN2a was trained using a 1-D tensor of the encoding epoch’s word-locked broadband iEEG recordings, that were labeled as encoded or forgotten based on the performance on the free recall block after that session’s distractor (Fig A3). CNN2b also classified words as encoded or forgotter but utilized the paired 1-D convolved HG and convolved beta tensors in the LSMG iEEG during encoding instead (Fig A3, 1B2).

To differentiate between good recall and poor recall, the LSMG iEEG during the entire 30 sec. free recall epoch was used to derive paired 1-D tensors of convolved HG and convolved beta Each session of the free recall epoch was assigned a binary value of good or poor recall based on whether the number of recalled words exceeded the mode of the number of recalled words across all the experimental sessions. The mode of the number of words recalled varied across the individual patients, with some subjects exhibiting a mode of 0 words recalled.

CNNs were implemented in Keras™ for binary classification. The CNN architecture used for all models consisted of pairs of Conv1D and Batch Normalization layers followed by a Global Average Pooling layer and a fully connected layer as the output layer. The CNN structure utilized for all CNNs was 3 convolutional 1-D layers (with 64, 128 and 64 units per layer), a Kernel size of 3, and an Adam optimizer. Hyperparameter tuning was performed by testing different structures (consisting of 2 or 3 convolutional layers with 32, 64, 128 or 256 units per layer) and choosing the one that maximized the area under the receiver operating curve (AUC) across all patients. Other hyperparameters were: 1) 100 epochs and a batch size of 128 for CNN1; 150 epochs and a batch size of 128 for analysis for CNN2a,b; and 100 epochs and a batch size of 32 for CNN3. For each patient, for CNN1,2a,and 2b the confusion matrices and related measures were derived using five-fold cross-validation across all the trials. The number of trials for all the CNNs in each fold varied by patient (Table 1-3), due to variability in the number of experimental sessions completed per patient.

**Table 1:** Confusion table metrics of encoding vs. recall trial classification using CNNs trained and cross-validated using paired one-dimensional LSMG convolved high-gamma and beta bursts, during the encoding and recall epochs (CNN1). Metrics calculated at Youden’s J of the receiver operating curve.

| Patient | accuracy | Auc | ppv | npv | sensitivity | specificity | f1 | sessions |
| --- | --- | --- | --- | --- | --- | --- | --- | --- |
| 1 | 0.992 | 1.000 | 1.000 | 0.985 | 0.985 | 1.000 | 0.992 | 40 |
| 2 | 0.698 | 0.743 | 0.712 | 0.712 | 0.692 | 0.705 | 0.691 | 24 |
| 3 | 0.754 | 0.849 | 0.744 | 0.756 | 0.683 | 0.825 | 0.698 | 10 |
| 4 | 0.660 | 0.698 | 0.670 | 0.637 | 0.438 | 0.882 | 0.502 | 24 |
| 5 | 0.628 | 0.625 | 0.649 | 0.659 | 0.640 | 0.614 | 0.611 | 32 |
| 6 | 0.698 | 0.733 | 0.798 | 0.699 | 0.611 | 0.796 | 0.639 | 8 |
| 7 | 0.933 | 0.965 | 0.937 | 0.932 | 0.930 | 0.937 | 0.933 | 50 |

In the case of CNN3 two-fold cross-validation was used because the sessions were relatively low in number (*i*.*e*., 12 trials per session). For individual patients the free-recall and categorical free-recall experiments were analyzed separately.

### Statistics

Mean values are accompanied by the standard error of the mean (s.e.m). A generalized linear model was used to predict the area under the receiver operating curve of CNN2b, for classifying successfully encoded words, using the two factors of consistent peak HG burst amplitude and consistent beta burst amplitude. The generalized linear model used a normal distribution and an identity link.

## Results

The data analyzed was generated during the Restoring Active Memory (RAM) project^1,11,12,22,23^. This project included hundreds of patients with medically-refractory focal epilepsy undergoing pre-surgical evaluation. Each of the patients performed the verbal delayed free recall task (Figure 1A). The multisite collaborative DARPA RAM research team examined whether the iEEG, from all the recording contacts and all spectra of brain activity during encoding, can be used as factors to train a logistic regression classifier to distinguish recalled words from forgotten words. The RAM team found that this method achieved a mean area under the receiver operating curve (AUROC) of 0.63 +/-0.07^1,11^. The logistic regression classifier utilized by the RAM studies is analogous to the CNN2a,b in our study (Figure 1A,1B2). Our approach was distinct from thar used in prior RAM projects^1,11^, because we: 1) utilized CNNs to classify both memory state and performance; 2) utilized iEEG recordings solely from the LSMG alone; and 3) we asked whether high-gamma (HG) and beta bursts are sufficient for defining verbal memory states (encoding vs. recall) and delayed free recall performance.

In 142 experiments, from 113 patients, we compared the AUROC of CNN2a and CNN2b for labeling recalled words during the encoding epoch (Figure 1A2,A3,1B2). CNN2a was trained and cross-validated using the 1-dimensional (1-D) tensor of broadband LSMG iEEG. CNN2b was trained and cross-validated using paired 1-D tensors of convolved HG and beta bursts derived from the LSMG iEEG. We found a strong linear correlation (R^2^=0.22, p<0.001) between the AUROC of CNN2a and CNN2b (Figure 4A). We next examined the subset of experiments in which CNN2b labeled recalled words better than chance (AUROC greater than 0.51, Figure 4A horizontal dashed line). Among this subset of 64 experiments from 64 subjects, we identified 14 patients, in which the CNN2b (i.e., LSMG iEEG convolved HG and beta) labeled recalled words with an AUROC greater than 0.6 (Figure 4B1, blue and green squares). We used an AUROC of 0.6 as a threshold since it is near the mean AUROC achieved in the prior RAM experiments^1,11^. In this subset of 14 patients, the AUROCs for labeling recalled words using CNN2a (*i*.*e*., broadband LSMG iEEG) were smaller than the equivalent AUROCs using CNN2b (HG and beta bursts) (Wilcoxon signed rank-test p=0.02, n=14,14, mean CNN2a AUROC=0.61+/-0.04, CNN2b AUROC=0.68+/-0.02 mean+/-s.e.m).

**Figure 4:**
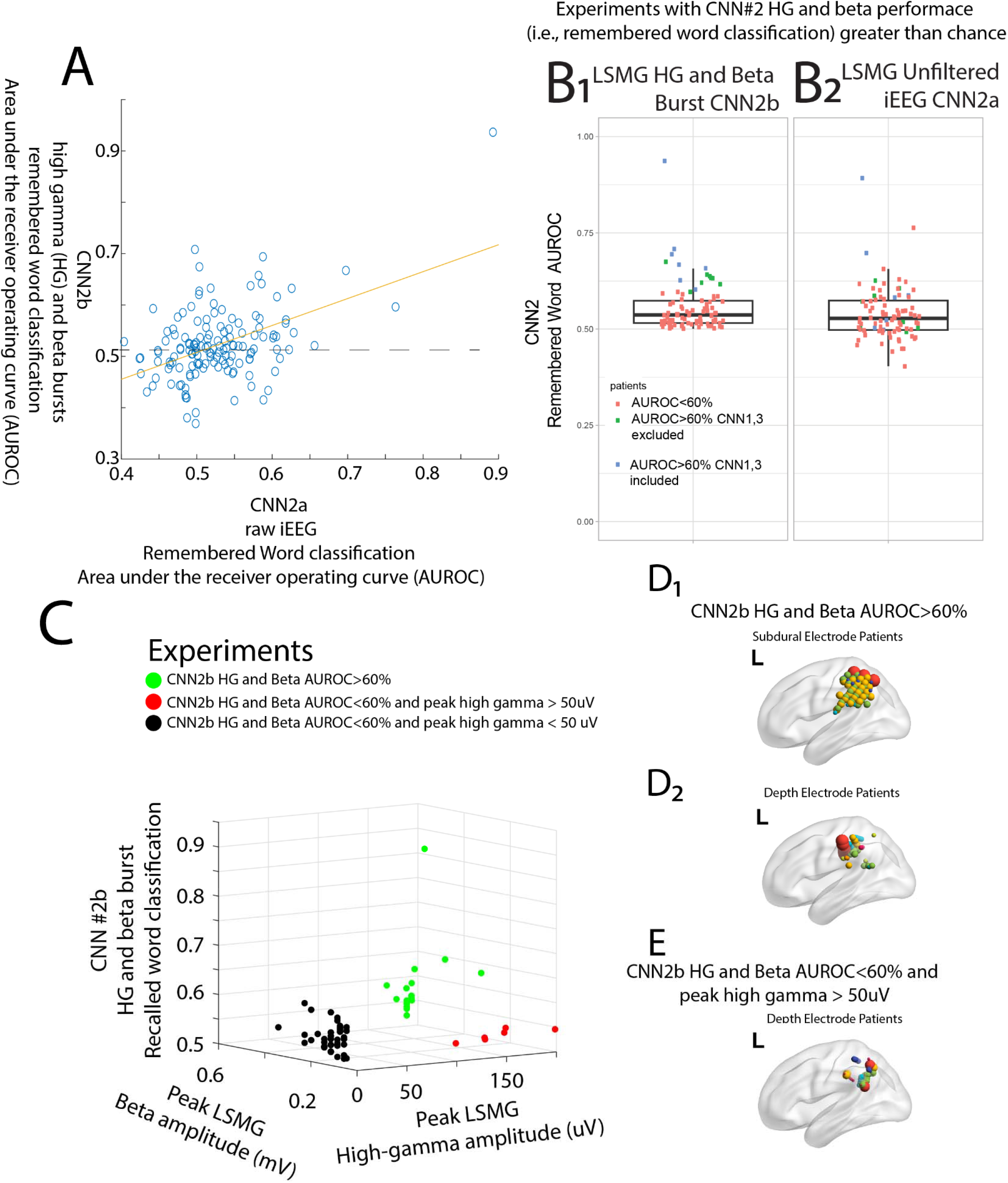
During word encoding, higher amplitude high-gamma (HG) and beta bursts in the anterior— dorsal region of the left supramarginal gyrus (LSMG) correspond with improved performance of the convolutional neural network 2b (CNN2b) in classifying recalled words. (A) Within-experiment comparisons of area under the receiver operating curve (AUROC) for classifying encoded words using CNNs that utilize 1-D tensors of the unfiltered LSMG iEEG (x-axis), or utilize paired 1-D tensors of LSMG convolved HG and beta (y-axis, CNN2b, R^2^=0.22, p<0.001). (B) The AUROC of CNN2b was greater than chance in 64 experiments (A, AUROC=0.52, horizontal dashed line). Among these 64 patients, the AUROC of CNN2b was greater than 0.6 in 14 subject—experiments (B1, blue and green squares). As shown in the box and scatter plots, in these 14 patients, the AUROC of CNN2a was less than CNN2b (B1,2, Wilcoxon signed rank-test p<0.05). (C) Among all the experiments shown in (B, n=64), a 3-D scatter plot compares the consistent peak HG and beta amplitudes for each experiment and each experiment’s corresponding CNN2b AUROC. Green circles indicate experiments with a CNN2b AUROC greater than 0.6 and consistent peak HG amplitude greater than 50uV (n=14), red circles patients with an CNN2b AUROC less than 0.6 but consistent peak HG amplitudes greater than 50 uV (n=6), and black circles a CNN2b AUROC less than 0.6 and consistent peak HG amplitude less than 50uV (n=44). (D) Location of electrode contacts for experiments with a CNN2b AUROC greater than 0.6 (D1:subdural contacts, D2:depth contacts). Colors indicate individual patients, and larger size contacts represent larger CNN2b AUROC values. (E) Location of depth electrodes for the patients in (C, red circles) with CNN2b AUROC less than 0.6 but HG amplitude greater than 50 uV. Note that in panel E the contacts are positioned more posterior—dorsal relative to panels D.

We next asked why CNN2b predicted recalled words with an AUROC greater than 0.6 in only 14 of the 64 patients (Figure 4B1, blue and green squares), and not in the other 50 experiments (Figure 4B1, red squares). We found that the LSMG iEEG during the encoding epoch exhibited consistent bursts of HG greater than 50uV in amplitude in all these 14 patients (Figure 4C, green circles). In 44 of the other 50 experiments the consistent HG bursts in the LSMG iEEG during the encoding epoch were less than 50uV in amplitude (Figure 4C, black circles). However, in 6 of the other 50 patients, consistent HG bursts greater than 50 uV were recorded from the LSMG iEEG during encoding but the CNN2b AUROC was less than 0.6 (Figure 4C, red circles). We utilized a generalized linear model and found that the consistent peak HG amplitude was significantly correlated with the CNN2b AUROC (d.f.=60, t=2.72, p=0.009), but peak beta amplitude (p>0.05) and the interaction of the peak HG and beta amplitude (p>0.05) did not reach significance.

We hypothesized that the location of depth electrode contacts in the LSMG may differ for the 14 patients with a CNN2b AUROC greater than 0.6 (Figure 4C, green circles) and experiments with a CNN2b AUROC less than 0.6, but consistent peak HG amplitudes greater than 50uV (Figure 4C, red circles). We found that the patients with a CNN2b AUROC greater than 0.6 had depth electrode contacts placed in the anterior-dorsal—most portion of the LSMG (Figure 4D2), whereas patients with a CNN2b AUROC less than 0.6 but exhibiting consistent HG amplitudes greater than 50uV had depth electrode contacts most posteriorly and ventral in the LSMG (Figure 4E). The experiments in patients with subdural electrode implants and a CNN2 AUROC greater than 0.6 had coverage of most of the LSMG (Figure 4D1), except for the most posterior-ventral regions (Figure 4E). We did not observe a linear correlation between the number of depth electrode contacts (n=102 experiments, t=1.93, p=0.05), or subdural grid and strip contacts (n=55 experiments, t=1.76, p=0.08) with a larger CNN2b AUROC.

In 7 of the patients with a CNN2b AUROC greater than 0.6 (Figure 4B1, blue squares) we performed pilot experiments to determine: 1) whether CNN1 (Figure 1A,1B1), trained and cross-validated using paired tensors of convolved HG and beta bursts from a randomized combination of labeled 3-second encoding epoch and recall trials could accurately label the encoding epochs; and 2) whether CNN3, (Figure 1A1,1B3) trained and cross-validated using paired tensors of convolved HG and beta bursts from the delayed free recall sessions, and dichotomously labeled based on whether the number of words recalled in a free recall session exceeded the mode, across sessions, would label the sessions in which correctly recalled words exceeded the mode; and 3) How would contingency table metrics differ across these seven patients for CNN1, CNN2b, and CNN3.

We found that in all seven patients the CNN1 (memory state: encoding vs. recall) AUROC was greater than 0.6, and in two of the patients the CNN1 was AUROC greater than 0.9 (Figure 1B1,Table 1). The F1 score for CNN1 ranged between [0.50-1.0]. The AUROC for CNN2b, which distinguished encoded from forgotten words during encoding, was more modest, but two patients exhibited a CNN2b AUROC greater than 0.7 (Figure 1B2, Table 2), the range of CNN2b AUROC was [0.6-0.9]. The F1 score for CNN2b ranged between [0.25-0.83]. Lastly, for CNN3 which distinguished good from poor recall, only two of the patients exhibited an AUROC greater than 0.6 (Figure 1B3, Table 3), and the CNN3 AUROC range was [0.44-0.86]. The CNN3 F1 score range was [.24-.87].

**Table 2:** Confusion table metrics of word encoded vs. word forgotten classification using CNNs trained and cross-validated using paired one-dimensional LSMG convolved high-gamma and beta bursts, during the stimulus locked encoding epoch. Metrics calculated at Youden’s J of the receiver operating curve. Words recalled are defined during the corresponding free recall epoch after the session’s distractor.

| Patient | Accuracy | Auc | ppv | npv | sensitivity | specificity | f1 | sessions |
| --- | --- | --- | --- | --- | --- | --- | --- | --- |
| 1 | 0.629 | 0.606 | 0.177 | 0.915 | 0.605 | 0.576 | 0.252 | 40 |
| 2 | 0.637 | 0.613 | 0.200 | 0.897 | 0.534 | 0.654 | 0.282 | 24 |
| 3 | 0.701 | 0.715 | 0.376 | 0.877 | 0.560 | 0.732 | 0.413 | 10 |
| 4 | 0.637 | 0.613 | 0.200 | 0.897 | 0.534 | 0.654 | 0.282 | 24 |
| 5 | 0.620 | 0.642 | 0.142 | 0.947 | 0.667 | 0.582 | 0.229 | 32 |
| 6 | 0.694 | 0.626 | 0.236 | 0.819 | 0.333 | 0.788 | 0.254 | 8 |
| 7 | 0.862 | 0.906 | 0.858 | 0.873 | 0.817 | 0.894 | 0.834 | 50 |

**Table 3:** Confusion table metrics of more words remembered vs. less words remembered using CNNs trained and cross-validated using paired one-dimensional LSMG convolved high-gamma and beta bursts, during the free recall epoch. Metrics calculated at Youden’s J of the receiver operating curve.

| Patient | accuracy | Auc | Ppv | Npv | sensitivity | specificity | f1 | sessions |
| --- | --- | --- | --- | --- | --- | --- | --- | --- |
| 1 | 0.567 | 0.575 | 0.208 | 0.658 | 0.300 | 0.700 | 0.243 | 40 |
| 2 | 0.840 | 0.860 | 0.947 | 0.693 | 0.828 | 0.871 | 0.876 | 24 |
| 3 | 0.613 | 0.440 | 0.162 | 0.730 | 0.260 | 0.746 | 0.196 | 10 |
| 4 | 0.557 | 0.479 | 0.351 | 0.643 | 0.400 | 0.658 | 0.329 | 24 |
| 5 | 0.520 | 0.508 | 0.350 | 0.517 | 0.317 | 0.733 | 0.313 | 32 |
| 6 | 0.583 | 0.543 | 0.494 | 0.606 | 0.530 | 0.626 | 0.487 | 8 |
| 7 | 0.724 | 0.790 | 0.728 | 0.775 | 0.709 | 0.736 | 0.683 | 50 |

## Discussion

Past attempts to derive biomarkers of memory performance during encoding of words for the verbal delayed free recall task have examined iEEG across all frequency spectra and among electrode contacts in diverse brain regions simultaneously^1,11^. This approach yields a mean AUROC of 0.62 for labeling successfully encoded words^1,11^. We found that in only 14 of the 113 patients, and 14 of the 142 experiments, that successful encoding of words could be predicted by the CNN2b with a similar accuracy. CNN2b was trained and cross-validated using paired tensors of convolved bursts of high-gamma (HG) and beta recorded solely from the left supramarginal gyrus (LSMG) during the word encoding epoch.

One factor that distinguished the 14 patients in our dataset with a CNN2b greater than 0.6 from the rest was that the CNN2b AUROC was significantly greater than the CNN2a AUROC. This result indicates that convolved high-gamma and beta bursts recorded from the LSMG were important in classifying encoding performance. A second factor distinguishing these 14 patients was consistent higher amplitude HG bursts recorded from depth or subdural electrodes in the LSMG. We did identify six other patients with higher amplitude HG bursts in the LSMG iEEG, but a CNN2b AUROC less than 0.6. Further investigation established that the locations of the electrode contacts were a third factor that distinguished the 14 patients. In the 6 patients exhibiting higher amplitude HG bursts but poor encoding classification performance, the location of the electrode contacts in the LSMG were distinct (posterior—dorsal) from the 14 other patients (anterior—ventral) with a CNN2b AUROC greater than 0.6.

Then, in pilot experiments in seven of the patients with a CNN2b AUROC greater than 0.6, we derived two other CNNs. CNN1 labeled verbal memory state (i.e., encoding versus recall) with an AUROC greater than 0.6 across all 7 subjects. CNN3 labeled better delayed free recall performance with an AUROC greater than 0.6 is two of the seven subjects. Both CNN1 and CNN3 utilized paired tensors of convolved bursts of high-gamma (HG) and beta recorded from the left supramarginal gyrus (LSMG). These results suggest that high-gamma and beta bursts in the LSMG are biomarkers of memory state and performance.

Prior work has shown the presence of subsequent memory effects (SME) in the LSMG using both fMRI^2,3,5^ and iEEG^9,10^. Delineating a SME relies on averaging responses to a stimulus and consequently, many SMEs coincide with stimulus presentation. HG and beta burst analyses have been utilized to better understand working memory and add additional information such as the onset, offset, frequency and duration of the burst ^18–20,24^. It has been proposed the gamma and beta bursts encode individual items in working memory^18–20,24^.

Therefore, burst analysis has some intrinsic advantages over examining trial averages and derivations of SME. In our approach, we did not utilize the spectral content of the discrete HG or beta. Future experiments can ask if using the specific spectral content of the HG and beta bursts in the convolved 1D tensors may improve the CNNs’ classification results.

Although we show that the CNNs can classify verbal episodic memory state and performance, this does not imply that, for example, more HG bursts or less beta bursts in the LSMG signify successful encoding of a memory engram or item ^18–20,24^. 1-D CNNs that utilize time series data^25^ are complex and difficult to interpret mechanistically^25^. This shortcoming is important because the mechanism used by the CNN to distinguish memory states and performance may be useful to support or refute competing mechanistic theory of LSMG function (i.e., attention to memory hypothesis (AtoM) versus the mnemonic accumulator hypothesis ^2–6^).

Even though CNNs are complex and difficult to interpret, CNNs are excellent for engineering applications. Also, two approaches that can be used to better understand the mechanism of 1-D CNNs are: 1) importance analysis that ranks the most important time series in the tensor with respect to classification; and 2) activity analyses which determine which specific epochs of the tensor that are most important to classification. We did not pursue importance analysis or activity analyses, and these techniques could have helped resolve mechanisms used by the four trained CNNs. For instance, CNN1 may have been biased by stimulus locked increases in HG that were present in the encoding trials but not the recall trials.

Lastly, we did not thoroughly investigate whether alternative machine learning strategies would be effective such as long short-term memory networks^26^, or temporal convolutional neural networks^27^ Further identification of the ideal machine learning strategy would ideally be carried out in experiments on patients that completed more experimental trials and sessions. There is also the possibility of using multiple machine learning methods and combining their results to enhance predictions^28^.

Understanding the neurophysiological correlates of verbal memory is better studied in epilepsy patients with intracranial electrodes as compared to scalp EEG due to decreased signal to noise, particularly in the high-gamma band ^29^. Despite this, some research shows that high-gamma (i.e., ripple band [80-200 Hz]) can be recorded from the scalp^30,31^. Utilizing sub-scalp electrodes improves the signal to noise ratio of detecting HG, ripple, and fast ripple (200-600 Hz) events^32^. Future research could examine whether sub-scalp electrode contacts^33^ over the LSMG can detect HG and beta bursts with a sufficient signal-to-noise ratio to train and test the CNNs described herein. It would also be of interest if these CNNs could predict performance in Weschler Memory Scale tasks and perhaps provide helpful biofeedback.

## Data Availability

All data produced in the present study are available upon reasonable request to the authors

https://openneuro.org/datasets/ds004789/versions/3.1.0

https://openneuro.org/datasets/ds004809/versions/2.2.0.

## Credit author statement

**Nicolás Sawczuk** - conceptualization, methodology, software, investigation, resources, writing – original draft, writing – review & editing. **Daniel Y. Rubinstein** software, investigation, resources, writing – original draft, writing – review & editing. **Michael R. Sperling -** investigation, writing – review & editing, funding acquisition. **Katrina Wendel-Mitoraj -** review & editing. **Petar Djuric** - review & editing. **Diego F. Slezak -** investigation, writing – review & editing, funding acquisition. **Juan Kamienkowski -** investigation, writing – review & editing, funding acquisition. **Shennan Aibel Weiss** - conceptualization, methodology, software, investigation, resources, writing – original draft, writing – review & editing

## Acknowledgements

The authors would like to thank Dr. Camarillo-Rodriguez, Mr. Zachary Waldman, Dr. Paul Wanda, Dr. Michael Kahana, Dr. Daniel Rizzuto, Dr. Sandhitsu Das, Dr. Daniel Rubinstein, Dr. Iren Orosz, Dr. Ashwini Sharan, Dr. Joel Stein, Dr. Bradley Lega, Dr. Robert Gross, Dr. Gregory Worrell, Dr. Katherine Davis, Dr. Barbara Jobst, Dr. Sameer Sheth.

## Data availability

Please see methods for links to the opensource dataset. The code used in this analysis is freely available upon reasonable request to the authors.

