## supplementary data for "High-gamma and beta bursts in the Left Supramarginal Gyrus can accurately differentiate verbal memory states and performance"

Health Sciences Tower

HSC T12, Room 041
Stony Brook, NY 11794-8121

Supplementary methods:

High-gamma (HG) and beta burst detection

The wavelet convolution was applied to the signals using the time_frequency.tfr.cwt function from the MNE Python library using complex Morlet wavelets. The frequency ranges were [5,50 Hz] for beta and [50,250 Hz] for HG (width=7 and gwidth=5). Within each window the iso-contours of power in the wavelet convolution were analyzed as described in (Waldman et al., 2018). The closed contour loop groups with an outermost contour that both surpassed a threshold of power (20% of the maximum power of the plot) and was within the desired frequency range (15-40 Hz for beta and 80-200 Hz for HG) were marked as oscillatory bursts. For each discrete beta and HG burst we defined the oscillatory burst power, frequency, and onset and offset times. For the encoding epoch, oscillatory bursts were detected with a sliding window of 0.2 seconds for HG and 0.6 seconds for beta. The sliding windows had an overlap of 0.1 seconds for HG and 0.3 seconds for beta. For the recall epoch, the topographical analysis of iEEG signals was performed similar to the encoding epoch but in overlapping 3 second segments(with 0.6 seconds of overlap) to generate a continuous HG and beta time series.
